# Focus or Feeding: A Quantitative Analysis of Prescribed Amphetamines in Human Milk

**DOI:** 10.64898/2026.10.14.26365136

**Authors:** Emily Yamada, Amy Stark, Jordan Burdine, Laxmi Donikini, Palika Datta, Kaytlin Krutsch

## Abstract

**Objective:** Amphetamine (AMP) and its derivates are commonly prescribed for the management of attention-deficit/hyperactivity disorder (ADHD). This study aimed to 1) quantify AMP’s transfer into human milk in lactating women receiving therapeutic doses and 2) contextualize infant exposure.

**Methods:** Milk samples and dyad health histories of thirteen lactating women treated with varying doses of either mixed AMP, dextroamphetamine, or lisdexamfetamine were released for this study from the InfantRisk Human Milk Biorepository. Milk drug concentrations were quantified by validated liquid chromatography-tandem mass spectrometry (LC-MS/MS) and used to calculate standard relative infant dose (RID_mat_) and RID standardized to the defined daily dose (RID_DDD_) of 15 mg AMP base.

**Results:** Average milk concentrations (C_avg_) of AMP base ranged from 23.3 to 297.0 ng/mL and increased linearly (R² = 0.92) with maternal daily dose. The RID_mat_ ranged 3.6–10.8% and RID_DDD_ ranged 1.4–21.8%. The fitted regression line crossed the 10% RID_DDD_ threshold at 1.17× DDD, with the upper 95% prediction limit crossing at 0.84× DDD. No adverse event reported by the mother–infant dyads were attributed to AMP exposure, notably including absence of reduced milk supply.

**Conclusions:** The levels of AMP observed in human milk followed a dose predictable manner. The average patient was predicted to remain below the 10% RID _DDD_ safety threshold at total daily doses at or below approximately 28 mg of mixed AMP salts or 59 mg of lisdexamfetamine. Ninety-five percent of patients were predicted to remain below the threshold at or below approximately 20 mg and 40 mg, respectively.

## Introduction

Amphetamine (AMP) stimulants approved by the U.S. Food and Drug Administration advise against breastfeeding, irrespective of salt form, formulation, or dose (1–3). Products carrying the old labeling format only state that AMP are excreted in human milk and mothers should refrain from nursing, without any supporting data (4). Products revised under the Pregnancy and Lactation Labeling Rule (PLLR) add quantitative pharmacokinetic detail, citing a relative infant dose (RID) of 2–13.8% and a milk-to-plasma ratio of 1.9–7.5; likewise they sustain the recommendation against breastfeeding (2). This recommendation rests on a narrow evidence base. Currently, published lactation pharmacokinetic literature for this class comprises of one case report, a cohort study of four women, and a retrospective neurodevelopmental follow-up of thirteen dyads in which milk was not sampled (5–7).

Pharmacokinetic evidence is vital when we consider the implications of women not continuing their attention-deficit/hyperactivity disorder (ADHD) therapy. Absent that evidence, and the cost of the recommendation falls on each individual patient. Women with moderate to severe ADHD who are also breastfeeding must either accept an uncharacterized infant exposure or discontinue treatment. This concern is particularly relevant given that approximately 60% of women on ADHD medications discontinue or interrupt treatment during pregnancy; moreover, discontinuation frequently persists into the postpartum period (8). Discontinuing breastfeeding is not necessarily a neutral alternative for mother or child. In addition to infant benefits, breastfeeding beyond 12 months also reduces maternal rates of hypertension, hyperlipidemia, and cardiovascular disease (9). Further, women with ADHD are roughly five times more likely to receive a postpartum psychiatric diagnosis than women without ADHD (10). These diagnoses may further exacerbate the challenges in early motherhood, including occupational disruption, executive dysfunction in parenting and household management, and overall increased maternal stress (11). These challenges compound postpartum sleep deprivation, which further compromises focus and executive function (11).

A recent review in the American Journal of Obstetrics & Gynecology recommends nonpharmacologic therapy for mild to moderate ADHD (11). For mothers with moderate to severe ADHD, stimulant use is considered acceptable, provided that clinicians use the lowest effective dose and the smallest number of medications possible (11). When stimulant therapy is continued during breastfeeding, the same review recommends close monitoring of the infant for potential adverse effects such as irritability, reduced weight gain, or poor sleeping patterns (11, 12). Applying that guidance requires knowing what infant exposure a given maternal dose produces.

Stimulants, including AMP and methylphenidate, remain the standard first-line pharmacologic therapies for ADHD (13). Methylphenidate is generally preferred during breastfeeding on the basis of lower milk transfer (12). AMP is currently available as a mixture of both the d- and l-isomer (Adderall; Adderall XR; Mydayis), the pure d-isomer dextroamphetamine (Dexedrine; ProCentra; Xelstrym; Zenzedi), and the d-isomer prodrug lisdexamfetamine dimesylate (LDX) (Vyvanse). Comparing AMP exposure across these formulations is complicated by the fact that labeled strengths are expressed as salt weight whereas pharmacokinetic assays quantify the free base. RID estimates calculated against labeled doses are therefore not directly comparable between these products (2).

RID carries a second limitation, independent of formulation: it is referenced to the mother’s own dose. Because milk concentration rises with dose, that denominator largely cancels the dose term, so women receiving substantially different doses can return similar RID values despite transferring different quantities of drug into milk.

Due to their mechanism of action, ADHD stimulants are classified as Schedule II controlled substances, reflecting a recognized potential for the development of substance use disorder. This understandably concerns mothers considering breastfeeding while taking these medications.

Currently, lactation references available to them and to their providers rest on the same limited primary data (14–16). This study addresses that gap by reporting the largest pharmacokinetic dataset to date on therapeutic-dose AMP transfer into human milk, standardized to AMP base equivalents, and referenced to the World Health Organization defined daily dose (DDD).

## Methods

Samples and data for this study were released by the InfantRisk Human Milk Biorepository (HMB) at Texas Tech University Health Sciences Center, Amarillo, under the Institutional Review Board approval (IRB No. A21-4214). All participants provided informed consent for the collection of human milk samples and associated maternal health data. Questionnaires collected infant age at sampling, diet, qualitative growth (weight gain, height growth, and head circumference), medical history, and any maternal-reported adverse events plausibly related to AMP exposure.

HMB samples were identified from participants who reported use of a prescribed AMP class medication in their medication questionnaire. Participants provided milk samples under steady-state conditions at predefined time points in their homes: 0, 1, 2, 4, 6, 8, 10, 12, 18, and 24 hours post-dose. One participant (#12), however, provided sampling spanning a single 6-hour dosing interval (timepoints 0–6 hours). Mothers were instructed that at each time point milk was to be expressed from both breasts, gently mixed to ensure homogeneity, and aliquoted into labeled collection tubes. Samples were frozen immediately following collection, shipped to the research facility under controlled conditions, and stored at −80°C until analysis.

Quantitative analysis was performed using an Agilent 1260 Series high-performance liquid chromatography (HPLC) coupled with an Agilent Ultivo mass spectrometer. Chromatographic separation was achieved using a Kinetex Biphenyl column (2.6 μm x 50 mm x 2.1 mm). The mobile phase consisted of (A) water with 0.1% formic acid and (B) acetonitrile with 0.1% formic acid, delivered at 0.3 mL/min using gradient elution. The injection volume was 5 μL. Mass spectrometric detection utilized electrospray ionization in positive ion mode under multiple reaction monitoring (MRM). The monitored transitions were m/z 136 → 91 for AMP and m/z 147 → 98 for the internal standard (AMP-d11).

The method was validated for selectivity, linearity (7.81–1000 ng/mL, r² > 0.99), precision, and accuracy, with sample preparation by protein precipitation. Full validation parameters are provided in the Supplementary Material. The analytical method measured total AMP.

More than half of FDA approved drugs are formulated as salts, and the strength printed on their label either refers to the salt or the pharmacologically active free base (17). The convention is set product-by-product rather than by any unified standard. Analytical assays quantify the free base, whereas prescribed doses are recorded in whichever unit the initial manufacturer label uses. Thus any calculation relating milk drug concentration to administered dose must reconcile the two or carry a systematic error. Published RID for this class have generally been calculated against labeled salt weight maternal doses without this reconciliation, which understates the RID (18).

Values reported here are not directly comparable to previously published estimates unless those are similarly corrected as well. Maternal daily doses were converted to AMP base equivalents using product specific factors. This was completed because the marketed salt forms are not interchangeable on a total salt weight basis: 0.626 for mixed d- and l-isomers, 0.733 for the pure d-isomer, and 0.297 for LDX (19, 20). To compare formulations via their AMP free base equivalents the following formulas were utilized (derivations, corroborating sources, and a worked example are provided in the Supplementary Material):

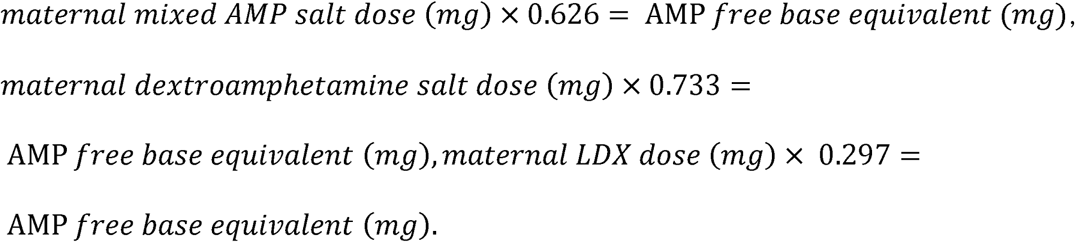

Once formulations were standardized, and the base equivalent dose summed across components for the single participant on a combination regimen, standard relative infant dose (RID_mat_) was calculated for the cohort (assuming 150 mL/kg/day milk intake, the standard value for an exclusively breastfed infant) via the following formula:

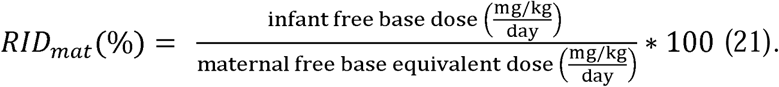

The RID formula with a maternal weight adjusted denominator has been used for decades in the absence of comparative infant doses; however, it carries a limitation across the wide maternal dosing range of AMP. With average milk concentration over the 24-hour dosing interval (C_avg_) being linear with the dose, the absolute infant dose also rises with maternal dose, while RID_mat_ remains constant, either over- or under-representing the individualized risk. To restore that contextualization, infant dose was additionally standardized against the World Health Organization (WHO) Defined Daily Dose (DDD) rather than maternal dose via the following

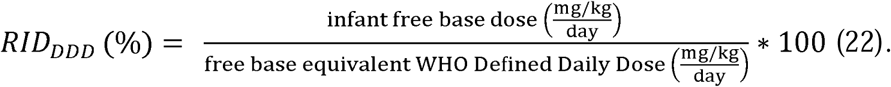

The World Health Organization Defined Daily Dose is 15 mg orally for AMP (N06BA01) and dextroamphetamine (N06BA02), and 30 mg for LDX (N06BA12) (23). These values are expressed as the active moiety (free base) rather than the salt, so no salt-to-base conversion was applied to the denominator. Supporting evidence is provided in the Supplementary Material.

Participants were subdivided by formulation category (immediate release [IR]; extended-release [ER]; repeated IR; IR + ER). Dose exposure relationships were modeled by ordinary least squares with fold-DDD as a continuous variable, with a 95% prediction interval used to identify the maternal dose below which 95% of new patients would remain below the 10% RID_DDD_ threshold.

## Results

Milk and data from thirteen participants were released from the HMB into the study (Table 1); the median maternal age was 34 years (range 28–39), while median weight was 75 kg (range 54– 99). Infant ages ranged from 1–3 months to 12–23 months, and all infants were reported to be receiving human milk for varying durations. The maternal formulation distribution was as follows: IR n = 2, repeated IR n = 2, IR + ER n = 1, ER n = 8. No participant reported underproduction of milk; eight reported producing about the right amount and five reported overproducing, with no apparent relationship to maternal dose. All five participants receiving at or above one DDD reported adequate or greater supply.

**Table 1.** Maternal characteristics and infant outcomes.

| # | Age | Race | Wt (kg) | Maternal Hx | Maternal Medication | Milk Supply | Infant Exposure | Infant Age | Infant Diet | Infant Growth (Wt/Ht/HC) | Infant Hx |
| --- | --- | --- | --- | --- | --- | --- | --- | --- | --- | --- | --- |
| 1 | 30-34 | white | 62 | ADHD | multivitamin, Vyvanse (lisdexamfetamine) (M-F) | Overproduce | BM: Y | 4-6 m | BM | above avg / above avg / above avg | reflux |
| 2 | 35-39 | white | 82 | ADHD, anxiety, asthma, migraines | Adderall (dextroamphetamine and amphetamine), alcohol (non-Rx) | Overproduce | BM: Y | 1-3 m | BM | above avg / above avg / — | dairy intolerance, frequent diarrhea, severe diaper rash |
| 3 | 35-39 | white | 70 | ADHD, anxiety, epilepsy, migraines | fluoxetine, lamotrigine, spironolactone-clindamycin, tretinoin, Vyvanse (lisdexamfetamine) | About right | BM: Y | 11-23 m | BM, water, solid | well above avg / avg / well above avg | — |
| 4 | 35-39 | white | 90 | ADHD, anxiety, asthma, bipolar disorder, chronic pain, depression, hypothyroidism, IBS, prolactinoma, sleep apnea | alcohol (non-Rx), atogepant, bupropion, cetirizine, choline, fluticasone propionate, multivitamin, Vyvanse (lisdexamfetamine) | Overproduce | BM: Y | 1-3 m | BM, formula | avg / avg / avg | — |
| 5 | 25-29 | white | 64 | ADHD, asthma, hyperlipidemia | Adderall (dextroamphetamine and amphetamine) XR, alcohol (non-Rx), multivitamin, tretinoin | About right | BM: 4 m | 7-11 m | BM, formula, solid | avg / avg / avg | — |
| 6 | 35-39 | white/asian | 86 | ADHD, asthma | alcohol (non-Rx), dextroamphetamine (Dexedrine), multivitamin, norethindrone | Overproduce | BM: 2 wk | 1-3 m | BM, formula | above avg / above avg / above avg | — |
| 7 | 30-34 | white | 67 | ADHD, anxiety, bipolar disorder, borderline personality disorder, depression | alprazolam, bupropion, fexofenadine, hydroxyzine, multivitamin, risperidone, topiramate, valacyclovir, Vyvanse (lisdexamfetamine) | About right | BM: Y | 7-11 m | BM, solid | avg / avg / avg | dairy intolerance, reflux, Trisomy 21 (congenital) |
| 8 | 30-34 | white | 67 | ADHD | Dextroamphetamine and amphetamine XR (Adderall XR), hydroxyzine, multivitamin | About right | BM: Y | 7-11 m | BM, water, solid | avg / avg / above avg | — |
| 9 | 30-34 | white | 74 | ADHD | Adderall (dextroamphetamine and amphetamine) XR, multivitamin | About right | BM: 7 wk | 1-3 m | BM | avg / avg / avg | — |
| 10 | 35-39 | white | 99 | ADHD, migraines | alcohol (non-Rx), desvenlafaxine XR, iron, melatonin, multivitamin, Mydayis (dextroamphetamine and amphetamine), THC (non-Rx) | Overproduce | BM: 6-10 m | 7-11 m | BM, water, rice cereal, solid | avg / avg / avg | hypoxic-ischemic encephalopathy (congenital) |
| 11 | 25-29 | white | 54 | ADHD, anxiety | Adderall (dextroamphetamine and amphetamine), fluoxetine, | About right | BM: 3 m | 7-11 m | BM, water, | below avg / avg / avg | frequent ear infection, |
|  |  |  |  |  | multivitamin |  |  |  | juice, rice cereal, solid |  | tongue/lip tie |
| 12 | 30-34 | white | 86 | ADHD, anxiety, asthma, congenital sucrase-isomaltase deficiency, GERD | Dextroamphetamine and amphetamine (Adderall), sertraline | About right | BM: 8 m | 12-23 m | BM, water, solid | avg / avg / avg | — |
| 13 | 30-34 | white | 73 | ADHD, anxiety, GERD, migraines, narcolepsy (type 1 with cataplexy) | acetaminophen, Adderall (dextroamphetamine and amphetamine), doxycycline hyclate, ibuprofen, metronidazole, Vyvanse (lisdexamfetamine) | About right | BM: 2 m | 1-3 m | BM | avg / avg / avg | — |

The milk concentration-time profiles are represented in Figure 1 and Table 2. Peak concentrations (C_max_) ranged from 64.4 to 598.0 ng/mL; time to peak (T_max_) was 1 hour for both IR participants and ranged from 1 to 8 hours among ER participants. C_avg_ ranged from 23.3 to 297.0 ng/mL (Figure 2 and Table 2). RID_mat_ across the cohort ranged 3.6–10.8%, as seen in Table 2. The RID_DDD_ ranged 1.4–21.8%, with two participants exceeding the 10% threshold (Figure 3 and Table 2).

**Figure 1.**
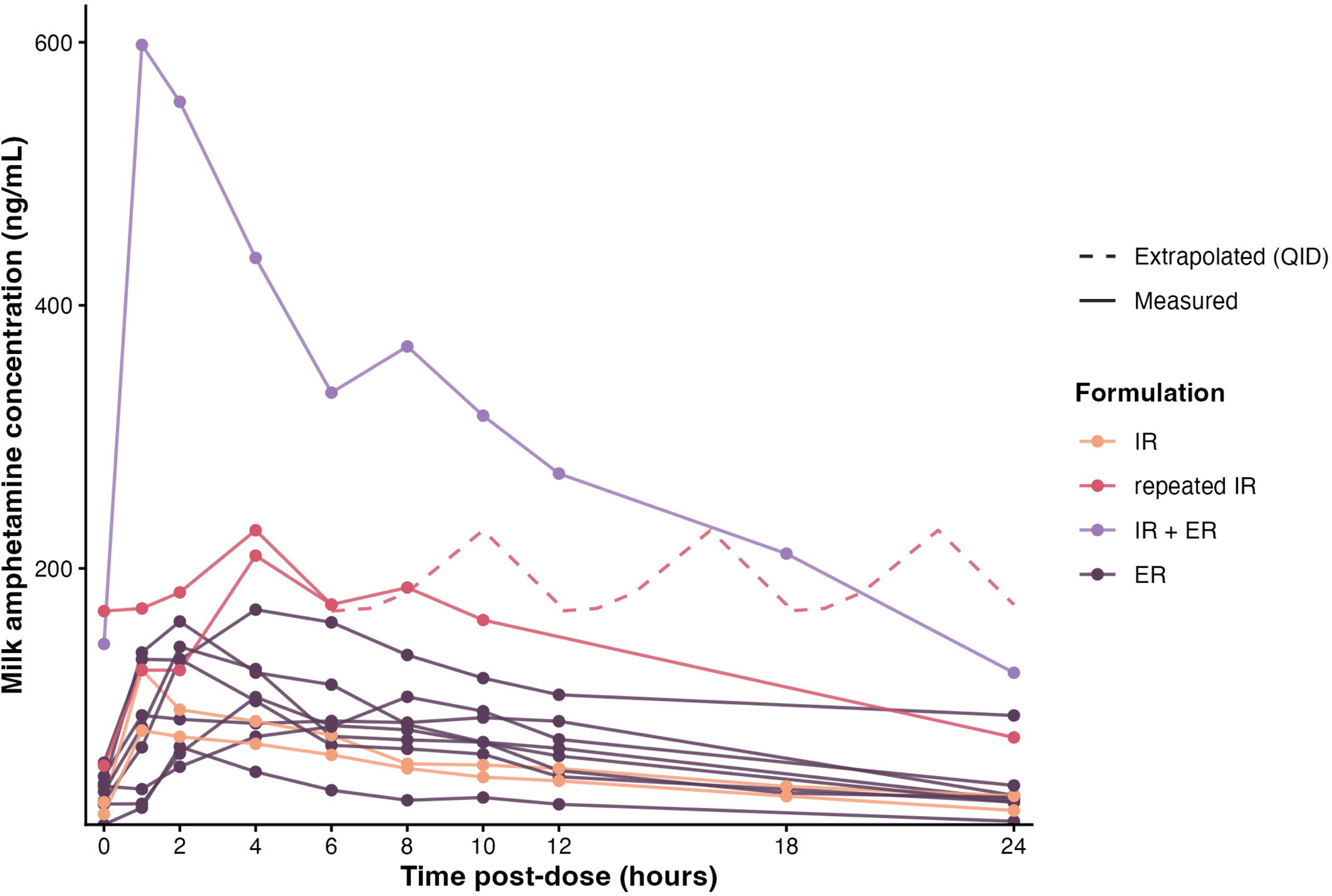
Milk AMP concentration (ng/mL) plotted against time post dose (hours) for all 13 analyzed participants, by formulation category (IR; repeated IR; IR + ER; ER). Each line represents a single participant; note that the y-axis scale differs between panels. LDX is grouped with ER products on the basis of duration of action rather than formulation technology; it is supplied as an IR capsule, and its duration derives from rate limited prodrug conversion. Participant #12 provided sampling across a single 6-hour dosing interval; her steady-state profile is shown replicated across the 24-hour day (dashed segments), and her AUC was extrapolated accordingly.

**Figure 2.**
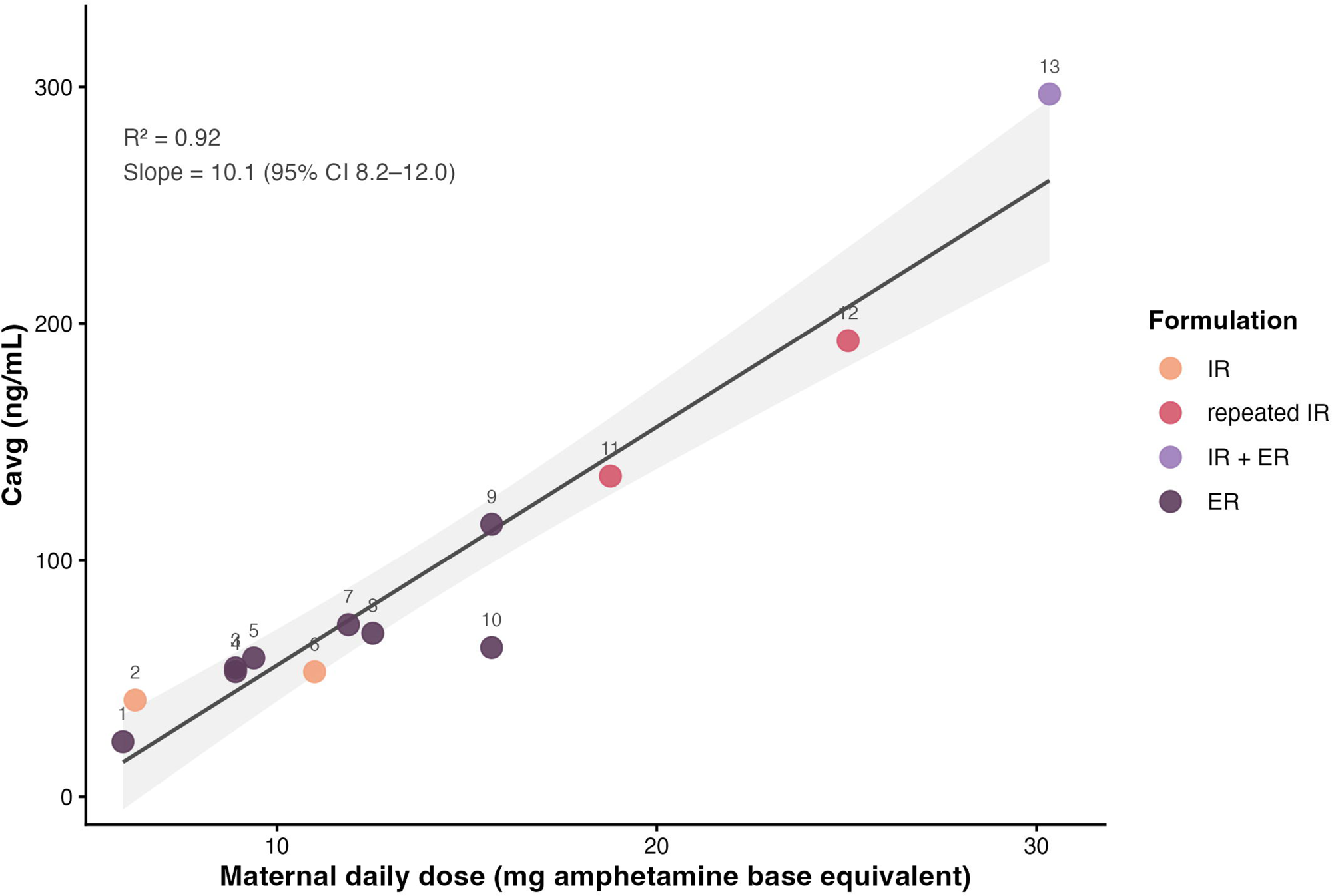
Average milk AMP concentration (C_avg_) against maternal daily dose in mg AMP base equivalents, with ordinary least squares regression. The maternal dose is expressed in mg AMP base equivalent using the product specific salt-to-base conversion factors. Points are labeled with the participant number used in Table 2.

**Figure 3.**
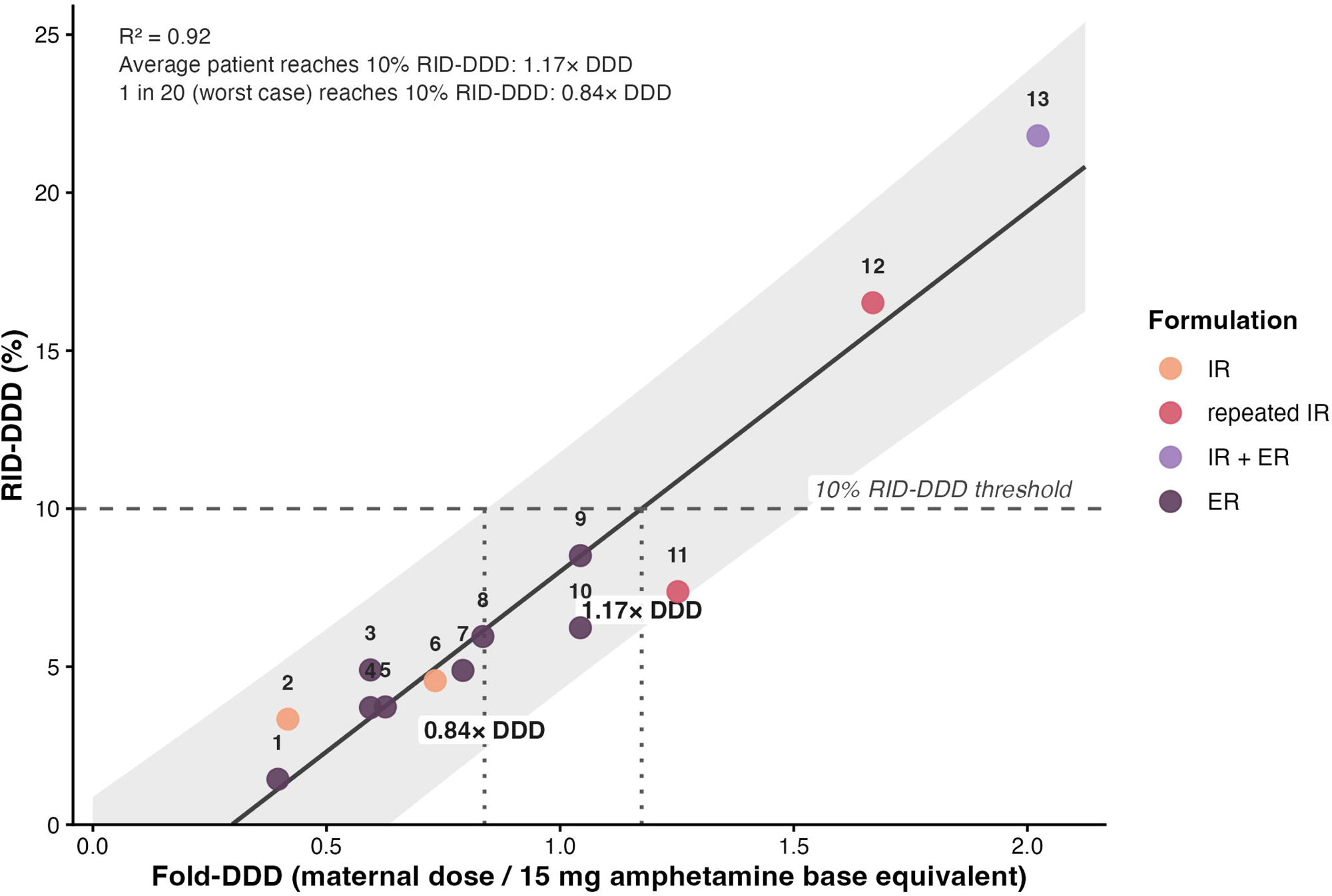
The solid line represents the ordinary least squares regression fit; the shaded band depicts the 95% prediction interval of where new individual patients from similar populations are expected to fall. The horizontal dashed line at 10% marks the conventional RID_DDD_ safety threshold.

**Table 2:** Per-patient pharmacokinetic results, ordered by ascending fold-DDD.

| # | Product | Daily dose<br>(mg base eq) | Fold-DDD | C<sub>max</sub><br>(ng/mL) | T<sub>max</sub><br>(h) | AUC<sub>0-24</sub><br>(ng*h/mL) | C<sub>avg</sub><br>(ng/mL) | RID<sub>mat</sub><br>(%) | RID<sub>DDD</sub><br>(%) |
| --- | --- | --- | --- | --- | --- | --- | --- | --- | --- |
| 1 | Vyvanse 20 mg daily | 5.9 | 0.4 | 64.4 | 2.0 | 560 | 23.3 | 3.6 | 1.4 |
| 2 | Adderall 10 mg daily | 6.3 | 0.4 | 76.7 | 1.0 | 982 | 40.9 | 8.0 | 3.3 |
| 3 | Vyvanse 30 mg daily | 8.9 | 0.6 | 102.1 | 4.0 | 1309 | 54.5 | 8.2 | 4.9 |
| 4 | Vyvanse 30 mg daily | 8.9 | 0.6 | 130.9 | 2.0 | 1270 | 52.9 | 6.2 | 3.7 |
| 5 | Adderall XR 15 mg daily | 9.4 | 0.6 | 140.3 | 2.0 | 1408 | 58.7 | 6.0 | 3.7 |
| 6 | Dexedrine 15 mg daily | 11.0 | 0.7 | 122.7 | 1.0 | 1268 | 52.9 | 6.2 | 4.6 |
| 7 | Vyvanse 40 mg daily | 11.9 | 0.8 | 159.7 | 2.0 | 1745 | 72.7 | 6.2 | 4.9 |
| 8 | Adderall XR 20 mg daily | 12.5 | 0.8 | 88.3 | 1.0 | 1659 | 69.1 | 7.1 | 6.0 |
| 9 | Adderall XR 25 mg daily | 15.7 | 1.0 | 168.6 | 4.0 | 2765 | 115.2 | 8.2 | 8.5 |
| 10 | Mydayis 25 mg daily | 15.7 | 1.0 | 102.3 | 8.0 | 1514 | 63.1 | 6.0 | 6.2 |
| 11 | Adderall 15 mg BID<br>(2<sup>nd</sup> dose 3 hours after 1<sup>st</sup>) | 18.8 | 1.3 | 209.7 | 4.0 | 3253 | 135.5 | 5.9 | 7.4 |
| 12 | Adderall 10 mg QID | 25.0 | 1.7 | 229.0 | 4.0 | 4625 | 192.7 | 9.9 | 16.5 |
| 13 | Vyvanse 60 mg + Adderall 10 mg BID | 30.3 | 2.0 | 598.0 | 1.0 | 7128 | 297.0 | 10.8 | 21.8 |

**Abbreviations**
**AUC<sub>0-24</sub>**: area under the curve for 24 hours
**BID**: twice a day
**C<sub>avg</sub>**: concentration average
**C<sub>max</sub>**: concentration maximum
**DDD**: daily defined dose
**QID**: four times a day
**RID<sub>DDD</sub>**: relative infant dose standardized to the defined daily dose
**RID<sub>mat</sub>**: standard relative infant dose
**T<sub>max</sub>**: time maximum

C_avg_ was noted to increase linearly (R² = 0.92) with maternal daily dose in AMP_base_ (Figure 2). And correspondingly, RID_DDD_ increased linearly with fold-DDD (Figure 3). The fitted regression line crossed the 10% RID_DDD_ threshold at 1.17× DDD, with the upper 95% prediction limit crossing at 0.84× DDD.

ER formulations (n = 8) accounted for the majority of the cohort. The combined IR + ER regimen (n = 1, #13) produced the highest absolute exposure; this single observation reflects the AMP base equivalent dose summed across both components (30.3 mg/day, 2.02× DDD).

Repeated IR dosing (n = 2) produced higher peak concentrations than single daily IR dosing; participant #12, who provided only 6 hours of sampling, had elevated trough concentrations consistent with accumulation from preceding doses (Figure 1) making our assumptions likely to overestimate true exposure.

Maternal doses ranged from 5.9 to 30.3 mg AMP base equivalent per day (Table 2), corresponding to 0.40× to 2.02× the base equivalent DDD. No participant receiving less than 1.5× DDD exceeded the 10% RID_DDD_ threshold, and both participants above it did (Figure 3).

## Discussion

Among the 13 lactating women treated with different AMP formulations for ADHD, milk transfer followed expected pharmacokinetic behavior. Infant exposure to AMP in milk is driven by the maternal dose and milk volume consumed. Although RID_mat_ calculations placed most infant AMP_base_ exposures below the conventional 10% RID threshold, RID_DDD_ described RID more usefully for clinical practice. At total daily doses at or below approximately 20 mg of mixed AMP salts or 40 mg of LDX, infant exposure remained below the conventional 10% RID safety threshold for 95% of patients. Above these doses, RID_DDD_ exceeded the threshold in a minority. The two participants receiving more than 1.5× DDD both exceeded the threshold, while no participant below that dose did. These estimates must be weighed against the well established maternal and infant benefits of breastfeeding, the risks of untreated or undertreated maternal ADHD, and the benefits of effective maternal drug therapy (24).

Current lactation literature reports maternal doses in salt based labeled doses. These doses require conversion before they can be compared with the quantity of active drug measured. This key distinction is essential when calculating RIDs, because RID calculations utilizing salt based maternal dosing use a larger denominator than free base equivalent drug detection or comparisons. The matched approach, with its smaller denominator, therefore produces higher RID estimates. The magnitude of this discrepancy is product specific. Labeled dose calculations understate RID_mat_ approximately 1.4× for dextroamphetamine sulfate, 1.6× for mixed AMP salts. Applying base equivalent conversion also tightened the observed dose exposure relationship in this cohort, with R² for C_avg_ against maternal dose rising from 0.76 to 0.92 (Supplementary Material). RID_DDD_ is unaffected by this distinction, because the WHO DDD is itself expressed as the active moiety.

AMP base equivalent standardization allowed maternal dose and RID estimates to be evaluated on a common clinical scale, enabling meaningful dose response interpretation across formulations. The WHO derived AMP base equivalent DDD was utilized as a reference dose comparator rather than each mother’s own dose. One DDD corresponds to approximately 24 mg daily of mixed AMP salts or 50 mg of LDX. For maternal doses below 0.84× DDD, the upper 95% prediction limit of regression remains below the 10% RID threshold. This suggests that fewer than 1 in 20 patients would be expected to exceed the 10% RID_DDD_ threshold at total daily doses of approximately 20 mg of mixed AMP salts or 40 mg of LDX. For maternal doses below 1.17× DDD (approximately 28 mg and 59 mg daily respectively), the average patient is expected to remain below the 10% RID_DDD_ threshold. This region between thresholds represents a zone in which interindividual variability becomes the dominant determinant of infant exposure. Although the average patient is expected to remain below the threshold, a minority may still exceed it.

These findings bear on how the labeling recommendation is constructed. The Lactation subsection created by the PLLR generally carries a risk summary stating that the developmental and health benefits of breastfeeding should be considered along with the mother’s clinical need for the drug and any potential adverse effects on the breastfed infant. Where infant exposure is unpredictable, a categorical recommendation may be warranted. Current AMP labeling instead adopts this “alternative” recommended PLLR construction, reserved for drugs whose potential for serious adverse reactions in the breastfed infant is judged to universally outweigh that benefit, and advises against breastfeeding without further assessment. The present data indicate that infant AMP exposure is not unpredictable: it rose linearly with maternal dose, and a dose range associated with exposure below the conventional threshold can be identified. On that basis, a categorical recommendation applied uniformly across the dose range constrains the ability to individualize treatment.

Although drug transfer was dose dependent, concentration-time profiles demonstrated substantial differences in the trough milk concentrations across the cohort. Participants receiving ER monotherapy at standard doses returned to near zero milk concentrations between daily doses.

This reflects clearance of the drug over each dosing interval. In contrast, participants receiving combined IR + ER regimens (#13) or repeated IR dosing (#12) maintained trough concentrations approaching or exceeding the peak concentrations seen in lower dose ER monotherapy participants. These differences between regimens have practical implications for feeding strategies. ER monotherapy produced relatively smooth concentration-time profiles, suggesting limited benefits from feeding timing modifications. For maternal doses that exceed the threshold dose, partial breastfeeding supplemented by formula or banked human milk may offer a viable approach to maintaining infant exposure within target ranges without requiring maternal treatment discontinuation. Infant exposure scales with milk volume as well as with milk concentration. An infant receiving complementary foods consumes proportionally less human milk than an exclusively breastfed neonate, and a nursing toddler less still. Therefore, the same maternal dose delivers a progressively smaller absolute infant dose at this point. The 150 mL/kg/day intake assumed throughout this analysis represents full breastfeeding and therefore an upper bound; for partially breastfed infants, these estimates overstate exposure in proportion to the fraction of the diet supplied by milk.

Despite several high RID_DDD_ values, no adverse event was attributed to AMP exposure in the dyad self-reported health questionnaire (Table 1). Reported infant adverse events included reflux, frequent ear infection/diarrhea, and severe diaper rash; however, none of the mothers reported associating these symptoms with the maternal therapy. These events are common in early infancy, and most infants were reported to have average or above-average growth. Two infants carried congenital diagnoses that predated and were unrelated to milk exposure. Further, labeling cautions that large AMP doses may reduce milk production, a concern extrapolated from early postpartum prolactin suppression without measured milk output, and maternal prolactin concentration may not govern production once lactation is established (2). No participant here reported underproduction.

As mentioned earlier, previous reports have described AMP transfer into human milk. One case report described AMP in human milk at a maternal dose of 20 mg daily, with no abnormalities noted in the infant at 2 years (5). A pilot study reported normal neurodevelopmental outcomes at 18-month median follow-up in 13 retrospectively surveyed mothers taking AMP and its derivatives, supporting compatibility from a developmental outcomes perspective (7). And another report of four mothers (15–45 mg daily) reported a median RID of 5.7% (range 3.9– 13.8%), calculated against salt weight maternal doses and with no conversion applied (6).

Recalculated on a base equivalent basis their median RID_mat_ is 7.7% (5.3–18.9%); standardized against the DDD, their median RID_DDD_ is 9.6% (3.9–21.9%) (6). The maximum RID_DDD_ in that cohort, 21.9%, is nearly identical to the maximum observed here, 21.8%, and both occurred in the participant receiving approximately twice the DDD (6). That cohort also illustrates the limitation motivating RID_DDD_. The woman receiving 45 mg daily returned an RID_mat_ of 10.0%, apparently at the conventional threshold, while her RID_DDD_ was 21.9%, the highest value in either cohort (6). The originally reported RID of 2–13.8% cited in every AMP product revised under the PLLR updated drug labels is drawn from these same two reports (2). Presented as a bare range, the historic RID of 2–13.8% reads as unexplained variability. The present data indicate otherwise. Both C_avg_ and RID_DDD_ rose linearly with maternal dose (R² = 0.92). High RIDs of AMP stimulants are a consequence of high maternal doses rather than of unpredictable variability.

Though methylphenidate is the preferred stimulant when breastfeeding, switching to methylphenidate is not a straightforward alternative. Stimulant response is idiosyncratic, with roughly 40% of patients responding equally to both classes, 28% preferentially to AMP, and 16% to methylphenidate (25). Thus, a forced class switch may trade a known effective regimen for an uncertain one. Furthermore, a Norwegian cohort study found that full breastfeeding for longer than six months was associated with fewer ADHD symptoms in children (26). Given findings like these, clinicians must also not discount the potential risks associated with withholding breastfeeding on the child.

This study has several limitations. The 0.84× DDD threshold derived from the upper 95% prediction interval is influenced by this cohort size and should be read as a statistically conservative estimate reflecting the sample size rather than as a fixed clinical dose ceiling. Long term infant outcomes were not assessed, and infant plasma drug concentrations were not measured. Although RID provides a practical estimate of infant exposure, future studies could strengthen these findings by measuring infant plasma concentrations directly; where clinically indicated, pediatricians can obtain plasma levels in breastfed infants to quantify systemic exposure individually, with concentrations exceeding 25% of the lower end of the adult therapeutic range generally regarded as unacceptable (21). One participant, on four times daily (QID) dosing, provided only a single six-hour dosing interval at steady state. She showed elevated baseline concentrations, indicating that sampling did not capture her first daily dose.

Thus, extrapolating the six-hour area under the curve (AUC) across the day likely overestimates her true exposure; the retained approach is a conservative upper bound, consistent with the study’s general tendency to avoid underestimating infant exposure. Finally, the analytical method measured total AMP without resolving the d- and l-isomers. Because the cohort included products ranging from mixed AMP salts (approximately 3:1 d:l) to LDX and dextroamphetamine (exclusively d-isomer), equal base equivalent doses do not necessarily represent equipotent exposures. Base equivalent standardization reflects mass rather than potency, which is the appropriate basis for estimating infant exposure, since exposure is determined by the quantity of drug transferred rather than by receptor activity.

Despite these limitations, therapeutic dose AMP demonstrated dose predictable transfer into human milk. These findings expand the existing pharmacokinetic literature and provide preliminary evidence to support shared decision making regarding continuation of AMP therapy for ADHD during breastfeeding at typical therapeutic doses, with individualized consideration at higher doses. For these dyads, the clinical question is not AMP versus human milk, but whether the therapy is appropriate for both members of the dyad.

## Conflicts of interest

Dr. Krutsch serves as Director of the InfantRisk Center of Excellence at Texas Tech University Health Sciences Center and is an author of *Hale’s Medications & Mothers’ Milk*. The InfantRisk Center conducts investigator-initiated and industry-sponsored research related to medication use during lactation. Dr. Krutsch has participated in consulting, advisory, and research activities involving pharmaceutical companies and research organizations related to lactation pharmacology. The authors declare that no external sponsor had any role in conception, design, analysis, interpretation, writing, or decision to publish this work. All other authors declare no conflict of interest.

## Funding

No direct funding.

## Institutional Review Board Statement

The InfantRisk Human Milk Biorepository is conducted in accordance with the Declaration of Helsinki and approved by the Institutional Review Board of Texas Tech University Health Sciences Center Amarillo (IRB no. A21-4214, first approved 22 February 2021). Deidentified data was released from the biorepository for this study.

## Informed Consent Statement

Informed consent was obtained from all subjects involved in the InfantRisk Human Milk Biorepository.

## Data availability

Data are available upon reasonable request to the corresponding author

## Acknowledgments

Conceptualization: K.K., P.D., and E.Y.; Data curation: P.D. and K.K.; Formal analysis: P.D. and K.K.; Funding acquisition: P.D. and K.K.; Investigation: P.D. and K.K.; Methodology: P.D. L.D and K.K.; Resources: P.D., L.D and K.K.; Validation: P.D. and L.D; Visualization: K.K., P.D., and E.Y.; Writing original draft: K.K., P.D., and E.Y..; Writing, review & editing: K.K., P.D., E.Y., A.S. and J.B. All authors have read and agreed to the published version of the manuscript.

During the preparation of this work, the authors used Claude Opus 4.5 (Anthropic) to assist with manuscript preparation and editing. All intellectual content, data analysis, interpretation, and scientific conclusions are the sole work of the authors, who take full responsibility for the accuracy and integrity of this work.

## Notes

### Author Declarations

The Institutional Review Board of Texas Tech University Health Sciences Center Amarillo gave ethical approval for the InfantRisk Human Milk Biorepository (IRB No. A21-4214; first approved February 22, 2021). Deidentified data from the biorepository were released for this study.

