## Supplemental Materials for "Focus or Feeding: A Quantitative Analysis of Prescribed Amphetamines in Human Milk"

**Supplementary Material**

**1. Salt-to-base conversion factors**

Prescribed doses of amphetamine (AMP) are recorded via the unit provided by the manufacture’s product label; for AMP products that unit is usually the salt. Analytical assays, on the other hand, quantify AMP as the free base, and the two are not interchangeable. Relative infant dose (RID) calculations that place a base derived numerator over a salt derived denominator understate infant exposure by the ratio of the two. The magnitude of the correction is product specific, and is dependent on the counterion rather than the isomer. Enantiomers are identical in mass, so the d:l ratio of a product has no bearing on its salt-to-base fraction.

Mixed AMP salt products (Adderall, Adderall XR, Mydayis) are comprised of four salts in equal parts by weight (1). Two of those carry counterions substantially heavier than half a sulfate, decreasing their weighted base fraction well below that of a pure sulfate salt:

| **Component salt** | **Stoichiometry** | **Base fraction** |
| --- | --- | --- |
| Dextroamphetamine saccharate | 2:1 | 0.563 |
| Amphetamine aspartate monohydrate | 1:1 | 0.472 |
| Dextroamphetamine sulfate | 2:1 | 0.734 |
| Amphetamine sulfate | 2:1 | 0.734 |
| Equal parts weighted average | — | 0.626 |

For each of the product classes, similar calculations were conducted to find the products factor. The following were the factors applied in the analysis:

| **Product class** | **Examples** | **Factor** |
| --- | --- | --- |
| Mixed amphetamine salts | Adderall, Adderall XR, Mydayis | 0.626 |
| Dextroamphetamine sulfate | Dexedrine, Zenzedi, ProCentra | 0.733 |
| Lisdexamfetamine dimesylate | Vyvanse | 0.297 |
| Already labeled as base | Dyanavel XR, Adzenys XR-ODT | 1.000 |

**2. Basis of the WHO defined daily dose**

The defined daily doses (DDD) in ATC group N06BA are 15 mg orally for AMP (N06BA01) and dexamphetamine (N06BA02), and 30 mg for lisdexamfetamine (LDX) (N06BA12) (2). These are expressed as the active moiety rather than the salt. The WHO ATC/DDD Index flags salt-based values in its Note field, and that field is empty for all three entries. No salt-to-base conversion was therefore applied to the denominator. A DDD is the assumed average maintenance dose for the main indication. Read as base, one DDD corresponds to approximately 24 mg/day of mixed AMP salts, 20 mg/day of dextroamphetamine, or 50 mg/day of LDX. Read as salt, the same entries would correspond to 15 mg and 30 mg daily, which are starting rather than maintenance doses (2).

**3. Worked example**

Participant 9, receiving 25 mg extended release mixed AMP salts once daily, body weight 73.9 kg. And for the single participant on a combination regimen (participant 13), base equivalent doses were summed across components before step 3: 60 mg LDX × 0.297 = 17.82 mg, plus 20 mg mixed AMP salts × 0.626 = 12.52 mg, giving 30.3 mg AMP base per day and a fold-DDD of 2.02×.

| **Step** | **Quantity** | **Value** |
| --- | --- | --- |
| 1 | Labeled maternal daily dose | 25 mg mixed amphetamine salts |
| 2 | × 0.626 → base equivalent | 15.65 mg amphetamine base/day |
| 3 | Weight adjusted maternal dose | 0.2118 mg/kg/day |
| 4 | AUC_0–24_ (linear trapezoidal) | 2,765 ng·h/mL |
| 5 | C_avg_ = AUC / 24 | 115.2 ng/mL |
| 6 | Daily infant dose = C_avg_ × 150 mL/kg/day | 0.01728 mg/kg/day |
| 7 | RID_mat_ = step 6 / step 3 | 8.16% |
| 8 | DDD 15 mg base, maternal weight adjusted | 0.2030 mg/kg/day |
| 9 | RID-DDD = step 6 / step 8 | 8.51% |
| 10 | Fold-DDD = step 2 / 15 mg | 1.04× |

**4. Recalculation of previously published estimates**

Ilett and colleagues reported a median relative infant dose of 5.7% (range 3.9–13.8%) in four women taking dexamphetamine sulfate (3). Their published values reproduce exactly from C_avg_ × 0.15 L/kg/day divided by the weight adjusted maternal dose, confirming that the denominator is the labeled salt dose and that no conversion was applied (3).

Individual body weights and labeled doses can be recovered from their published weight adjusted doses: 58 kg on 15 mg, 70 kg on 45 mg, 67 kg on 15 mg, and 73 kg on 20 mg, permitting recalculation on all three bases (3).

| **Measure** | **Ilett et al. (n = 4)** | **Present cohort (n = 13)** |
| --- | --- | --- |
| RID_orig_, as published by WHO | 5.7% (3.9–13.8) | 3.7% (1.1–6.2) |
| RID_mat_, base equivalent maternal dose | 7.7% (5.3–18.9) | 6.2% (3.6–10.8) |
| RID_DDD_, against 15 mg base | 9.6% (3.9–21.9) | 4.9% (1.4–21.8) |

*Median (range). Whether Ilett's milk concentrations represent free base or salt is not stated; their analytical standard was dextroamphetamine sulfate. The base corrected rows assume free base.*

The three measures rank the two cohorts differently. On RID_mat_ the earlier cohort appears to reach substantially higher exposure (18.9% against 10.8%), driven by their patient C, who had unusually high milk transfer on a low dose. Dividing by a small maternal dose inflates RID_mat_ regardless of the absolute quantity the infant receives. The RID_DDD_ of the two cohorts converge almost exactly at the upper end, 21.9% against 21.8%, because both contain one participant at approximately twice the DDD.

Their patient B illustrates the limitation of RID_mat_ directly. At 45 mg daily she returns an RID_mat_ of 10.0%, apparently at the conventional threshold, while her RID_DDD_ is 21.9%. The large maternal denominator conceals an absolute infant exposure more than twice that of a woman at one DDD. This is the same behavior described in the Methods. Due to milk concentration rising linearly with dose, RID_mat_ remains approximately constant while absolute infant dose does not.

**5. Comparison of the three relative infant dose calculations**

Three calculations are compared. RID_orig_ is the conventional approach used in the published lactation literature for this class. RID_mat_ applies the same maternal dose denominator after conversion to AMP base equivalents. RID_DDD_ replaces the maternal dose with the WHO DDD of 15 mg AMP base. Milk concentrations, C_max_, T_max_, and AUC are identical across all three.

|  | **RID_orig_** | **RID_mat_** | **RID_DDD_** |
| --- | --- | --- | --- |
| Denominator | Labeled salt dose | Base equivalent dose | 15 mg base (DDD) |
| Median | 3.7% | 6.2% | 4.9% |
| Range | 1.1–6.2% | 3.6–10.8% | 1.4–21.8% |
| Participants above 10% | 0 of 13 | 1 of 13 | 2 of 13 |
| R² against maternal dose | 0.000 | 0.483 | 0.924 |
| p | 0.98 | 0.008 | <0.001 |

*R² for RID_orig_ and RID_mat_ is against their own dose denominator; for RID_DDD_ it is against fold-DDD.*

The final two rows are the substantive result. RID_orig_ bears no relationship to maternal dose in this cohort (slope −0.001, R² = 0.000, p = 0.98). A woman on 80 mg and a woman on 10 mg resolve as indistinguishable values. This is not a failure of the data but an arithmetic property of the measure. Because milk concentration rises approximately linearly with dose, dividing infant dose by maternal dose cancels the dose term. Thus, the resulting ratio is uninformative about how much drug the infant actually receives. RID_DDD_, which holds the denominator fixed, recovers the dose–exposure relationship almost completely (R² = 0.924).

RID_mat_ falls between the two (R² = 0.483) rather than being perfectly flat. This is because the fitted C_avg_ dose relationship has a negative intercept (−45.1 ng/mL); the ratio of concentration to dose therefore rises modestly across the observed dose range. The direction is the same as for RID_orig_. The correction from RID_orig_ to RID_mat_ is product specific, because it is governed by the fraction of the labeled weight that is AMP base:

| **Product class** | **RID_orig_ understates RID_mat_ by** |
| --- | --- |
| Lisdexamfetamine dimesylate (prodrug) | 3.4× |
| Combination lisdexamfetamine + mixed salts | 2.6× |
| Mixed amphetamine salts | 1.6× |
| Dextroamphetamine sulfate | 1.4× |

Three consequences follow. First, every RID_orig_ value is understated, whether or not more than one product is involved. The error is a direct multiple of the conversion factor, so a LDX study reporting an RID_orig_ of 3.0% is in fact reporting 10.1%. In the present cohort, participant 13 returns an RID_orig_ of 4.1% against an RID_mat_ of 10.8%, above it. Recalculating the four participants reported by Ilett and colleagues moves one of them from 7.3% to 10.0%, likewise across the threshold. Thus, threshold based conclusions drawn from RID_orig_ are not reliable. Second, RID_orig_ values are not comparable across products. Two women receiving identical AMP base, one as LDX and one as dextroamphetamine sulfate, differ approximately 2.5-fold in RID_orig_ purely as an artifact of labeling convention. Any table or review that places uncorrected values from different AMP products side by side is comparing quantities on different scales. Lastly, the correction improves model fit in a multi-product cohort. R² for C_avg_ against maternal dose rises from 0.76 to 0.92 here. This effect is specific to cohorts spanning products with different conversion factors. In a single product study, the conversion rescales every denominator by the same constant. Thus, the fitted slope changes but R² does not; therefore the reported values are still wrong, but the internal dose–response is undisturbed.

Unfortunately, fold-DDD is affected in the same way. Using labeled doses directly against the 15 mg DDD would place a participant taking 20 mg LDX at 1.33× DDD rather than 0.40×. The combination regimen participant at 5.33× rather than 2.02×, and would weaken the RID_DDD_ relationship from R² = 0.924 to 0.719.
